# Genetically proxied inhibition of interleukin-6 signaling: opposing associations with susceptibility to COVID-19 and pneumonia

**DOI:** 10.1101/2020.09.15.20165886

**Authors:** Susanna C. Larsson, Stephen Burgess, Dipender Gill

## Abstract

The inflammatory cytokine interleukin-6 (IL-6) is pivotal for orchestrating the immune response. Inhibitors of IL-6 signaling are being investigated as treatments for severe coronavirus disease 2019 (COVID-19). We conducted a Mendelian randomization study investigating the effect of IL-6 signaling on susceptibility to COVID-19 and pneumonia. Our results showed that genetically proxied inhibition of IL-6 signaling was associated with reduced risk of COVID-19, but also with increased risk of pneumonia. Respiratory disease is a main feature of severe COVID-19, and the potential of IL-6 signaling inhibitors to increase risk of pneumonia warrants vigilance and caution in their application to treat COVID-19.

---

The inflammatory cytokine interleukin-6 (IL-6) is central to orchestrating the immune system. The pathophysiological process underlying severe coronavirus disease 2019 (COVID-19), caused by the severe acute respiratory syndrome coronavirus 2, consists of an exaggerated host immune response and elevated circulating levels of inflammatory cytokines, including IL-6.^1^ As such, immunomodulatory agents are being investigated for the treatment of COVID-19. Glucocorticoids may limit inflammation-mediated lung injury in patients with severe COVID-19, and consequently reduce progression to respiratory failure and death. The RECOVERY trial found that administration of dexamethasone resulted in lower 28-day mortality among hospitalized COVID-19 patients who were receiving either invasive mechanical ventilation or oxygen alone at randomization, but not among those who were not receiving any respiratory support.^2^ Inhibition of IL-6 signaling may represent another potential immunomodulatory strategy for treating COVID-19,^3^ and a recent meta‐analysis of mean IL‐6 concentrations demonstrated 2.9‐fold higher levels in patients with complicated COVID‐19 compared with patients with non-complicated disease.^4^

Genetic variants that proxy inhibition of IL-6 signaling may be used as instrumental variables in the Mendelian randomization paradigm to investigate corresponding drug effects. Here, we conducted a Mendelian randomization investigation to assess the potential effect of pharmacological inhibition of IL-6 signaling on susceptibility to COVID-19 and pneumonia.

Inhibition of IL-6 signaling was proxied using seven single-nucleotide polymorphisms within or adjacent to the *IL6R* gene region that were associated with C-reactive protein (CRP; downstream molecule of IL-6 signaling) levels at the genome-wide significance threshold in 204 402 individuals of European ancestry.^5^ These genetic variants also had associations with fibrinogen, IL-6 and soluble IL-6 receptor in a pattern consistent with their effect on reducing IL-6 signaling.^5^

Summary-level genetic data for COVID-19 were acquired from: 1) The COVID-19 Host Genetics Initiative genome-wide association meta-analysis (round 3),^6^ which included 6696 COVID-19 cases vs. 1 073 072 population controls, 3199 hospitalized COVID-19 cases vs. 897 488 population controls, and 928 hospitalized COVID-19 cases vs. 2028 non-hospitalized COVID-19 controls; and 2) a genome-wide association study involving 1610 hospitalized patients with severe COVID-19, defined as having respiratory failure and requiring some degree of respiratory support, and 2205 control participants (healthy volunteers, blood donors and outpatients of gastroenterology departments) from seven hospitals in Italy and Spain.^7^ Corresponding data for pneumonia were obtained from: 1) the FinnGen consortium, including 15 778 cases of all pneumoniae (International Statistical Classification of Diseases and Related Health Problems – 10th Revision codes: J12-J16, J18), and 119 867 control participants;^8^ and 2) the UK Biobank cohort (through the MRC-IEU consortium via the MR-Base platform^9^), which included 6572 self-reported cases of pneumonia (id:UKB-b:4533) and 456 361 control participants. The genetic association estimates in all studies were adjusted for sex and ancestry. Only summary-level data were analyzed in this study, for which appropriate ethical approval and participant consent had previously been obtained.

We performed summary data Mendelian randomization analysis using the inverse-variance weighted method with multiplicative random-effects and accounting for correlations between genetic variants. Our results showed that genetically proxied inhibition of IL-6 signaling, scaled per 0.1 standard deviation lower CRP level, was associated with a reduced risk of COVID-19, but also with an increased risk of pneumonia (Figure 1).

**Figure 1.**
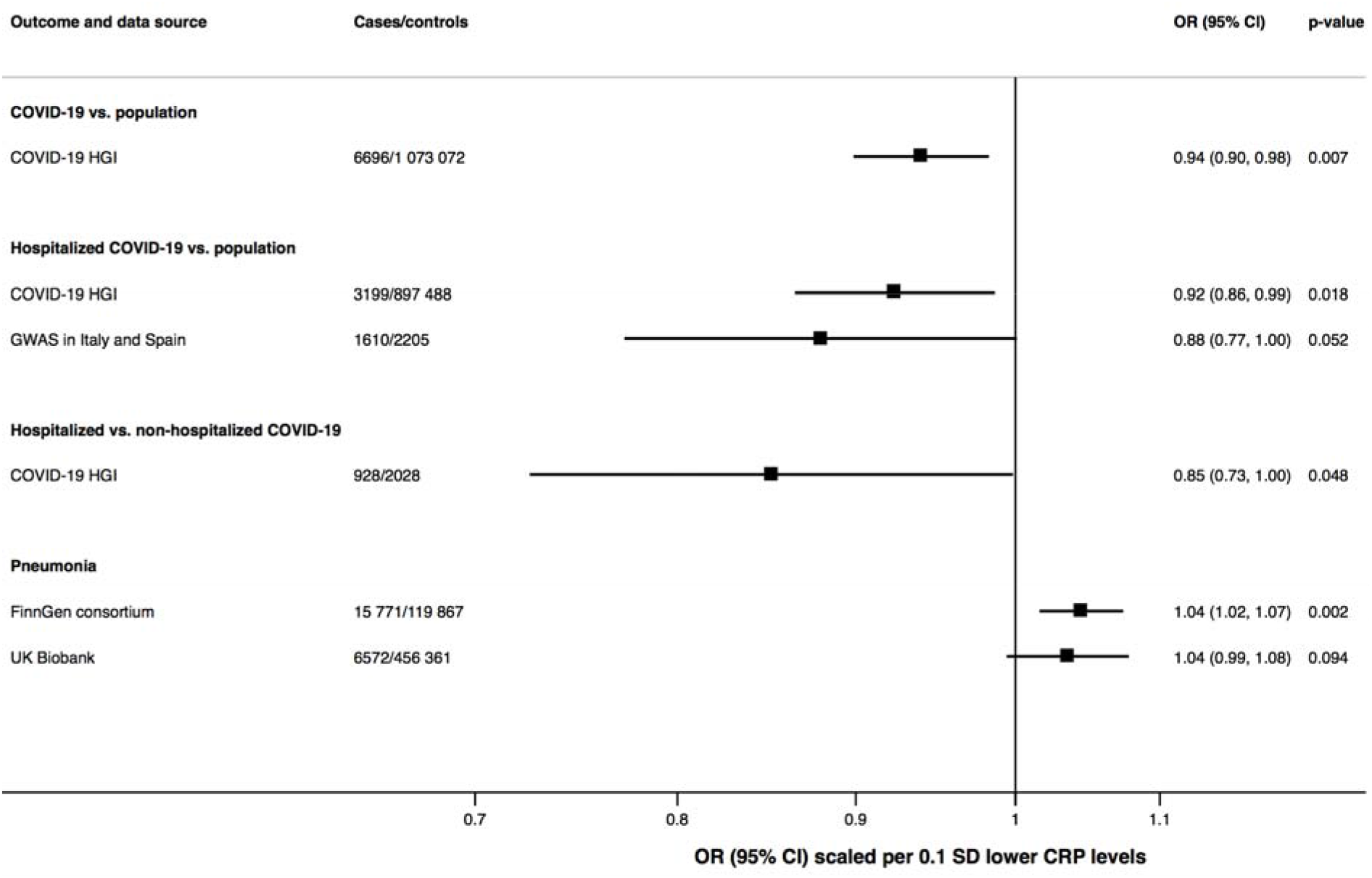
Associations of genetically proxied inhibition of IL-6 signaling with susceptibility to to COVID-19 and pneumonia. Estimates were derived using the multiplicative random-effects inverse sevariance weighted method and accounting for the correlations between the seven genetic variants in in or near the IL6 gene region. CI, confidence interval; CRP, C-reactive protein; GWAS, genome-wide association study; HGI, Host Genetics Initiative; OR, odds ratio; SD, standard deviation.

Our findings provide evidence supporting that inhibition of IL-6 signaling reduces susceptibility to COVID-19. Importantly, they go further to provide genetic evidence that inhibition of IL-6 signaling may also increase susceptibility to pneumonia. While we considered a broad definition of pneumonia from any cause, our results may be of relevance to secondary lung infections that can complicate COVID-19. An adverse effect of IL-6 inhibition on risk of pneumonia risk is biologically plausible, and respiratory tract infections, including pneumonia, are a well-known complication of IL-6 signaling inhibition.^10^ Respiratory disease is a main feature of severe COVID-19, and the potential of IL-6 signaling inhibitors to increase risk of pneumonia warrants vigilance and caution in their application to treat COVID-19.

## Data Availability

This study is based on publicly available summary-level data.

## Acknowledgments

The authors thank the investigators of the COVID-19 genome-wide association study, the COVID-19 Host Genetics Initiative, and FinnGen consortium for sharing summary-level data.

## Contributors

SCL, SB and DG contributed to the study design. SCL conducted statistical analyses. SCL and DG wrote the initial draft of the manuscript. All authors participated in the data interpretation and contributed to the final draft of the manuscript with intellectual importance.

